# Machine Learning for Mechanical Ventilation Control

**DOI:** 10.1101/2021.02.26.21252524

**Authors:** Daniel Suo, Cyril Zhang, Paula Gradu, Udaya Ghai, Xinyi Chen, Edgar Minasyan, Naman Agarwal, Karan Singh, Julienne LaChance, Tom Zajdel, Manuel Schottdorf, Daniel Cohen, Elad Hazan

## Abstract

We consider the problem of controlling an invasive mechanical ventilator for pressure-controlled ventilation: a controller must let air in and out of a sedated patient’s lungs according to a trajectory of airway pressures specified by a clinician.

Hand-tuned PID controllers and similar variants have comprised the industry standard for decades, yet can behave poorly by over- or under-shooting their target or oscillating rapidly.

We consider a data-driven machine learning approach: First, we train a simulator based on data we collect from an artificial lung. Then, we train deep neural network controllers on these simulators. We show that our controllers are able to track target pressure waveforms significantly better than PID controllers.

We further show that a learned controller generalizes across lungs with varying characteristics much more readily than PID controllers do.

## 1 Introduction

Invasive mechanical ventilation is a common medical treatment required for applications spanning anaesthesia, neonatal intensive care, and life support for the current COVID-19 pandemic.

Despite its importance, the core technology of medical ventilation has remained largely unchanged for years. It is common for a clinician to continuously monitor and adjust the ventilator for a patient manually.

Our work centers on the question of whether machine learning (ML) can give rise to control methods that are more robust, safer, and require less manual intervention. Specifically, we examine an ML-based methodology where we learn the dynamics of the ventilator-lung system and use the resulting differentiable simulators to train controllers. This isolation of tasks helps us better understand ventilator-lung behavior and allows us to more effectively train superior controllers.

We also study how our learned controllers and classical controllers are able generalize across lungs with varying physical characteristics. Such capability is crucial to automating more of the ventilator so it can adapt to patient needs, save clinicians’ time, and enable non-expert users to operate.

However, it is difficult to evaluate new control paradigms using commercial ventilators as they are closed systems both in terms of hardware and software. Thus, to demonstrate the potential advantage of machine learning for ventilator control, we validate our approach on a modular, low-cost, and fully open-source ventilator (LaChance et al., 2020) designed in response to the COVID-19 pandemic.

### Challenges of ventilator control

The main challenges of control in any clinical setting are robustness, safety, and the high cost of exploration. The controller must adapt quickly and reliably across the spectrum of clinical conditions, which are only indirectly observable. This is the reason that a simple and robust controller, namely the classic three-parameter Proportional-Integral-Derivative (PID) controller, is the method of choice. Few parameters require few samples to tune, and generalize more robustly to variations in the lung’s physical parameters.

To go beyond simple models and towards controllers based on machine learning models, we require significantly more data, which is impractical to collect on a human lung. Learning a reliable simulator is the problem of *nonlinear system identification*, which can be a statistically ill-posed problem in the worst case: there are unstable policies for which the closed-loop dynamics exhibit chaos. We study a residual approach to the safe exploration of the system’s dynamics.

We also consider the challenge of generalizing across dynamical systems. While the PID controller has few parameters and thus does exhibit some generalization, its performance on lungs with physical characteristics it was not tuned for is in general unsatisfactory, and fine-tuning or manual intervention by a clinician is required.

### 1.1 Our contributions

#### Improving over industry-standard baselines

We demonstrate the potential of an end-to-end pipeline for learning a controller, improving upon PID controllers for tracking pressure waveforms. These improvements are measured based upon ISO standards for performance of ventilatory support equipment (ISO 80601-2-80:2018). Our solution is composed of *“real2sim”* (learned simulator) and *“sim2real”* (learned controller on the learned simulator) components.

#### Improving generalization across lung dynamics

We demonstrate that a learned controller can improve performance simultaneously across several different ISO lung parameters. Our single controller improves upon *the best PID tuned for individual lung parameters*.

#### Model-based control with a learned physical model

Many recent works, under the research program of sample-efficient reinforcement learning, are concerned with long-term planning under a learned model of the environment’s dynamics. Several recent influential works learn a differentiable simulation of an environment that is itself simulated (like MuJoCo physics, or the rules of chess). Our work learns a differentiable simulation from physical data. We discuss the additional challenges that arise.

#### Simulators for ventilator dynamics

We provide a self-contained differentiable simulated environment for the ventilation problem, ready to be benchmarked with more sophisticated algorithms or modified to create more challenging settings (like zero-shot learning, or spontaneous perturbations). This reduces the entrance cost for future researchers to contribute to invasive mechanical ventilation. We emphasize that these settings must be properly addressed before this pipeline is suitable for clinical use.

### 1.2 Structure of the paper

In the next subsection 1.3 we discuss relevant related work. In Section 2 we present the necessary background on both control and ventilation physics. We also touch upon the motivation for our model-based approach. We describe our hardware setup for the mechanical ventilator in Section 3. The details on how we learn and evaluate a data-driven differentiable simulator for the mechanical ventilator are provided in Section 4. In Section 5 we describe the learning procedure for our controller and report on the final experimental results of its evaluation. Additional details on the simulator and controller learning are provided in the Appendix.

### 1.3 Related work

The modern positive-pressure ICU mechanical ventilator has history dating back to the 1940’s. For a comprehensive survey of the history of mechanical ventilation and its immense benefit to medical practice see Kacmarek (2011). The recent COVID-19 pandemic has prompted the publication of several open-source ventilator designs (Bouteloup et al., 2020).

Physics based models for ventilation are described in Marini & Crooke (1993). For a survey of control methods for ventilation see Chatburn & Mireles-Cabodevila (2011). Recent work has focused on augmenting the PID controller with adaptive control methods (Hazarika & Swarup, 2020; Shi et al., 2020). To the best of our knowledge, our learned differentiable simulator approach coupled with a trained deep model for the controller is novel in this field.

#### Control and RL with a learned model

Much progress has been made on end-to-end learned dynamics when the dynamics themselves exist *in silico*: MuJoCo physics (Hafner et al., 2019), Atari games (Kaiser et al., 2019), and board games (Schrittwieser et al., 2020). Combining such data-driven models with either pixel-space or latent-space planning has been shown to be effective for policy learning. A manifestation of this research program for the current deep learning era is established in Ha & Schmidhuber (2018).

#### Control and RL in physical systems

Progress on deploying end-to-end learned agents (i.e. controllers) in the physical world is more limited in comparison, due to difficulties in scaling parallel data collection, and higher variability in real-world data. Bellemare et al. (2020) present a case study on autonomous balloon navigation, adopting a Q-learning approach, rather than model-based like ours. Akkaya et al. (2019) use domain randomization in simulation to close the sim2real gap for a difficult dexterous manipulation task.

#### Multi-task reinforcement learning

Part of our methodology has close parallels in multi-task reinforcement learning (Taylor & Stone, 2009), where the objective is to learn a policy that performs well across diverse environments. During *global* training, we optimize our policy simultaneously on a learned ensemble of models corresponding to different physical settings, similar to the work of Rajeswaran et al. (2016); Chebotar et al. (2019) on robotic manipulation.

#### Machine learning for health applications

Healthcare offers a multitude of opportunities and challenges for machine learning; for a recent survey, see (Ghassemi et al., 2020). Specifically, reinforcement learning and control have found numerous applications (Yu et al., 2020a), and recently for weaning patients off mechanical ventilators (Prasad et al., 2017; Yu et al., 2019, 2020b). As far as we know, there is no prior work on improving the control of ventilators using machine learning.

## 2 Scientific background

### 2.1 Control of dynamical systems

We begin with some formalisms of the control problem. A fully-observable discrete-time dynamical system is given by the following equation:

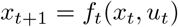

Here *x*_*t*_ is the state of the system, *u*_*t*_ is the control input and *f*_*t*_ is the transition function. Given a dynamical system, the control problem is to minimize a sum of cost functions over a long-term horizon:

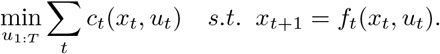

This problem is in general computationally intractable, and theoretical guarantees are available for special cases of dynamics (notably linear dynamics) and perturbations. For an in-depth exposition on the subject, see the textbooks by Bertsekas (2017); Zhou et al. (1996); Tedrake (2020).

#### PID control

An extremely ubiquitous technique for the control of dynamical systems is the use of linear error-feedback controllers, i.e. policies that choose a control based on a linear function of the current and past errors vs. a target state. That is,

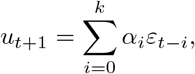

where 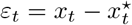 is the deviation from the target state at time *t*, and *k* represents the history length of the controller. PID applies a linear control with *proportional, integral*, and *differential* coefficients, i.e.

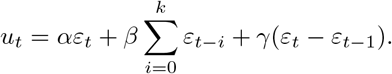

This special class of linear error-feedback controllers, motivated by physical laws, is known to perform and generalize well (Åström & Hägglund, 1995; Bennett, 1993). It is currently the industry standard for ventilator control.

### 2.2 The physics of ventilation

In invasive ventilation, a patient is connected to the ventilator with a hollow tube that goes into their mouth and down into their main airway or trachea. The ventilator is connected to a central pressure chamber, and applies pressure in a cyclic manner to simulate healthy breathing. During the inspiratory phase, the target applied pressure increases to the peak inspiratory pressure (PIP). During the expiratory phase, the target decreases to the positive-end expiratory pressure (PEEP), maintained in order to prevent the lungs from collapsing. The PIP and PEEP values, along with the durations of these phases, define the time-varying target *waveform*, specified by the clinician.

The goal of ventilator control is to regulate the pressure sensor measurements to follow the target waveform 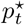. As a dynamical system, we can write *p*_*t*+1_ = *f*_*t*_(*p*_*t*_, *u*_*t*_), and define the cost function to be a measure of the deviation from the target; e.g. the absolute deviation 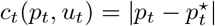. The objective is to design a controller that minimizes the total cost over *T* time steps.

A ventilator needs to take into account the structure of the lung to determine the optimal pressure to induce. Such structural factors include *compliance* (*C*), or the change in lung volume per unit pressure, and *resistance* (*R*), or the change in pressure per unit flow.

In designing and manufacturing ventilators, a standard proxy for the patient’s lung is a mechanical *test lung*, a physical instrument with calibrated bellows and springs which attach to the ventilator’s tubing. We use the hardware setup from the open-source People’s Ventilator Project (LaChance et al., 2020), which is outlined in Section 3.

#### Physics-based model

A simplistic formalization of the ventilator-lung dynamical system can be derived from the physics of a connected two-balloon system, with a *latent state v*_*t*_ representing the volume of air inside the lung. The dynamics equations can be written as

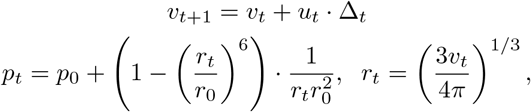

where *p*_*t*_ is the measured pressure, *v*_*t*_ is volume, *r*_*t*_ is radius of the lung, *u*_*t*_ is the input air flow rate, and Δ_*t*_ is the time increment. *u*_*t*_ originates from a pressure difference between lung-pressure *p*_*t*_ and supply-pressure *p*_supply_, regulated by a valve: 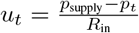. The resistance of the valve is *R*_in_ α 1*/d*^4^ (Poiseuille’s law) where *d*, the opening of the valve, is controlled by a motor, which is the control signal. The constants *p*_0_, *r*_0_ depend on both the lung and ventilator.

### 2.3 Challenges and benefits of a model-based approach

The first-principles dynamics model is highly idealized, and is suitable only to provide coarse predictions for the behaviors of very simple controllers. We list some of the various sources of error arising from using physical equations for model-based control:

- *Idealization of physics:* oversimplifying the complexity of fluid flow and turbulence via ideal incompressible gas assumptions; linearizing the dynamics of the lung and ventilator components.
- *Lagged and partial observations:* assuming instantaneous changes to volume and pressure across the system. In reality, there are non-negligible propagation times for pressure impulses, delayed pressure feedback arising from lung elasticity, and computational latency.
- *Underspecification of variability:* different patients’ clinical scenarios, captured by the latent proportionality constants *p*_0_, *r*_0_, may intrinsically vary in more complex (i.e. higher-dimensional) ways.

On the other hand, it is highly desirable to adopt a model-based approach in this setting, for its sample-efficiency and reusability. A reliable simulator enables the much cheaper and faster collection of data for training a controller, an ability to incorporate multitask objectives and domain randomization (e.g. different waveforms, or even different patients). An additional goal is to make the simulator *differentiable*, enabling direct gradient-based policy optimization through the system’s dynamics (rather than stochastic estimates thereof).

We show that in this partially-observed (but single-input single-output) system, we can query a reasonable amount of training data in real time from the test lung, and use it offline to learn a differentiable simulator of its dynamics (*“real2sim”*). Then, we complete the pipeline by leveraging interactive access to this simulator to train a controller (*“sim2real”*). We demonstrate that this pipeline is sufficiently robust that the learned controllers can outperform PID controllers tuned directly on the test lung.

## 3 Experimental Setup

To develop simulators and control algorithms, we run mechanical ventilation tasks on a physical test lung (IngMar, 2020) using the open-source ventilator designed by Princeton University’s People’s Ventilator Project (PVP) (LaChance et al., 2020).

### 3.1 Physical test lung

For our experiments, we use the commercially-available adult test lung, “QuickLung”, manufactured by IngMar Medical. The lung has three lung compliance settings (*C* = *{*10, 20, 50*}* mL/cmH2O) and three airway resistance settings (*R* = *{*5, 20, 50*}* cmH2O/L/s) for a total of 9 settings, which are specified by the ISO standard for ventilatory support equipment (ISO 80601-2-80:2018). An operator can change the lung’s compliance by engaging one or more springs that modify the difficulty of inflating the lung. To change airway resistance, the operator changes the diameter of the inlet to restrict or relax airflow. We connect the test lung to the ventilator via a respiratory circuit (McIntyre, 1986; Parmley et al., 1972) as shown in Figure 1.

**Figure 1:**
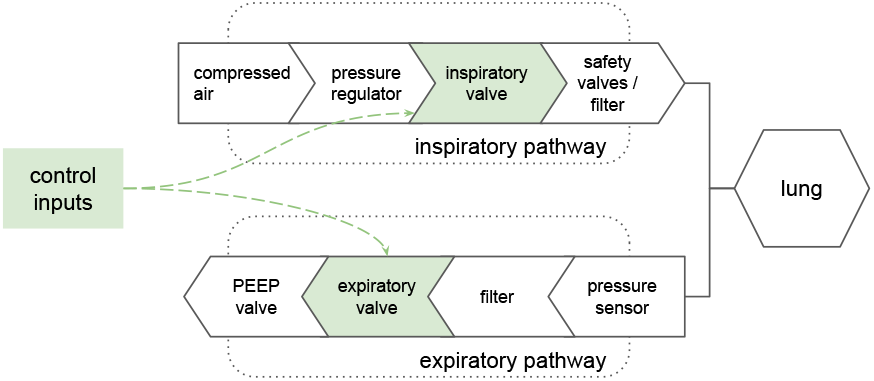
A simplified respiratory circuit showing the airflow through the inspiratory pathway, into and out of the lung, and out the expiratory pathway. We shade the components that our algorithms can control in green.

### 3.2 Mechanical ventilator

There are many forms of ventilator treatment. In addition to various target trajectories, clinicians may want to choose the physical quantity being targeted (e.g., volume, flow, or pressure of air), what ends a breath, and whether or not the patient initiates any breaths (Chatburn, 2007).

The PVP ventilator focuses on targeting pressure for a completely sedated patient (i.e., the patient does not initiate any breaths) and comprises two pathways (see Figure 1): (1) the inspiratory pathway that directs, regulates, and monitors airflow into the lung and (2) the expiratory pathway that does the same for airflow out of the lung. A software controller is able to adjust one valve for each pathway. The inspiratory valve is a proportional control flow valve that allows control in a continuous range from fully closed to fully open. The expiratory valve is a binary on-off valve that only permits zero or maximum airflow.

To prevent damage to the ventilator and/or injury to the operator, we implement four software overrides that abort a given run by closing the inspiratory valve and opening the expiratory valve: 1) if pressure in the lung exceeds 70 cmH2O, 2) if volume in the lung exceeds 1.2L, 3) if pressure in the lung drops below PEEP (5 cmH2O) for more than 300ms (indicates disconnected tubing), or 4) if we observe that the average sampling period exceeds 40ms over 300ms (indicates software delays). The PVP pneumatic design also includes a safety valve, which lets out air whenever internal pressure is in excess of 1 PSI (70 cmH2O), in case software overrides fail.

### 3.3 Abstraction of the ventilation task

We treat the mechanical ventilation task as episodic by extracting each inspiratory phase (e.g., light gray regions in Figure 2) from all breath timeseries as individual episodes. This approach reflects both physical and medical realities. Mechanically ventilated breaths are by their nature highly regular and feature long expiratory phases (dark gray regions in Figure 2) that end with the ventilator-lung system close to its initial state. What is more, the inspiratory phase is both most relevant to clinical treatment and most suboptimal for PID controllers (e.g., under- or over-shooting, ringing). Episodes of inspiratory phases are thus simplified, faithful units of training data for learning a differentiable simulator.

**Figure 2:**
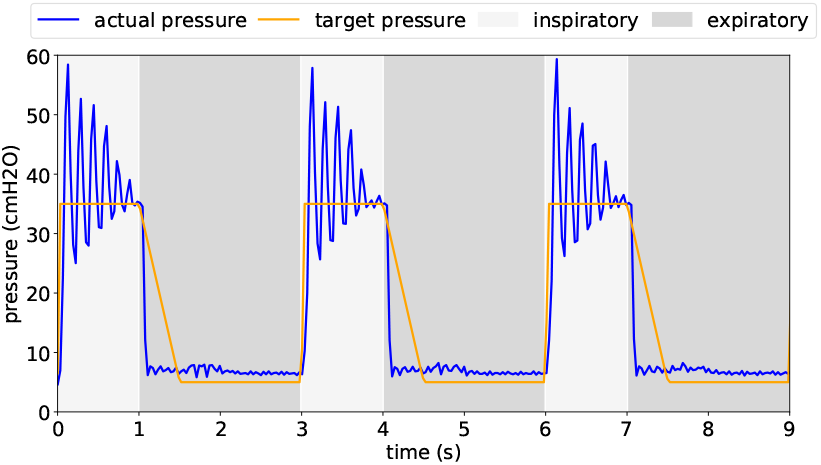
PID controllers exhibit their suboptimal behavior (under- or over-shooting, ringing) primarily during the inspiratory phase. Note that we use a hard-coded controller during expiratory phases to ensure safety. This choice does not affect our results.

## 4 Learning a data-driven simulator

With the hardware setup outlined in Section 3, we have a physical dynamical system suitable for benchmarking, in place of a true patient’s lung. In this section, we present our approach to learning a simulated surrogate of this ventilator-lung system, subject to the practical constraints of real-time data collection.

Two main considerations drove our simulator design:

- Evaluation of our simulator can only be done in a **black-box metric**, since we do not have explicit access to the dynamics equation, and existing physical models are poor approximations to the empirical behavior.
- The dynamical system we simulate is too complex for a comprehensive simulation and exhibits chaotic behavior in boundary cases. We can only hope to simulate it for “reasonable” scenarios. We thus explore within a neighborhood of realistic open loop trajectories.

### 4.1 Black-box simulator evaluation metrics

To evaluate our data-driven simulator for the mechanical ventilation system, we take into consideration that

1. Access to the physical system is black-box only and costly in terms of time/compute. That is, we have no explicit approximation of the dynamics equation.
2. The simulator dynamics are complex non-linear operations given by a deep neural network.

For these reasons we deviate from standard distance metrics considered in the literature, such as Ferns et al. (2005), as they explicitly involve the value function over states, transition probabilities or other unknown quantities. Rather, we consider metrics that are based on the evolution of the dynamics, as studied in Vishwanathan et al. (2007).

However, unlike the latter work, we take into account the particular distribution over policies that we expect. We thus define the following distances between dynamical systems. Let *f*_1_, *f*_2_ be two dynamical systems over the same state-action spaces. Let *µ* be a distribution over policies mapping state to action *π ∈* Π. Let *D* be a distribution over sequences of controls denoted *{u*_1_, *u*_2_, …, *u*_*T*_ *}*.

1. We let the **open-loop distance** w.r.t. horizon *T* and control sequence distribution *D* be defined as

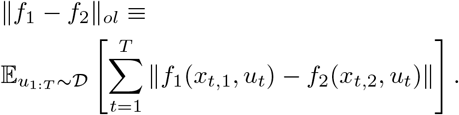
2. We let the **closed-loop distance** w.r.t. horizon *T* and policy distribution *µ* be defined as

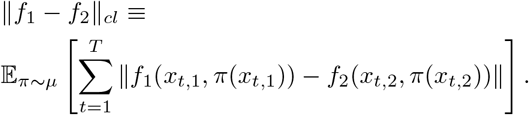

In both cases the inner norm is Euclidean over the states, although this can be generalized to any metric. We evaluate our data-driven simulator using the open loop distance metric, which we illustrate in the top half of Figure 3. In the bottom half, we show a sample trajectory of our simulator and the ground truth. See Section 4.3 for experimental details.

**Figure 3:**
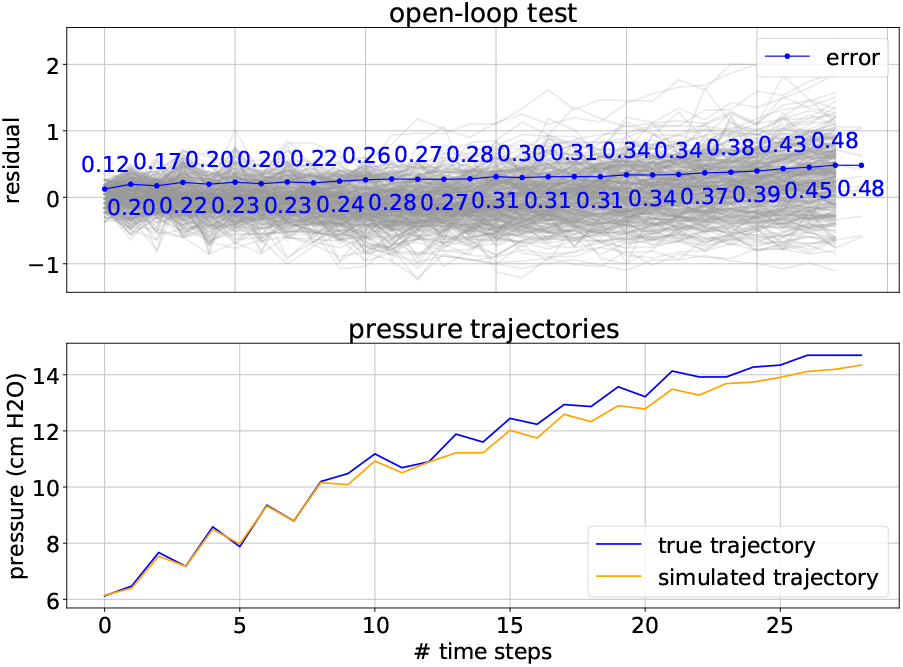
We show our simulator performance. The upper plot shows the *open-loop distance* and the lower shows a sample trajectory with a fixed sequence of controls (open-loop controls), both for lung setting *R* = 5, *C* = 50. In the former, we see low error as we increase the number of steps we project and in the latter, we see that our simulated trajectory tracks the true trajectory quite closely.

### 4.2 Targeted exploration

Due to safety and complexity issues, we cannot exhaustively explore the ventilator-lung system. Instead, we explore around policies generated by a baseline PI controller. For each of the lung settings, we collect data according to the following protocol:

1. Choose a safe PI controller baseline.
2. Introduce random exploratory perturbations according to the following two policies:
  a. Boundary exploration: to the very beginning of the inhalation, add an additional control sampled uniformly from 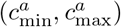 and decrease this additive control linearly to zero over a time frame sampledrandomly from 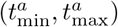;
  b. Triangular exploration: sample a maximal additional control from a range 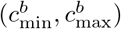 and an interval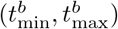, within the inhalation. Start from 0 additional control at time 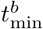, increase the additional control linearly until 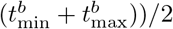, and then decrease to 0 linearly until 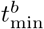.
  For each breath during data collection, we choose policy (*a*) with probability *p*_*a*_ and policy (*b*) with probability (1 *− p*_*a*_). The ranges in (*a*) and (*b*) are lung-specific. We give the exact values used in the Appendix.

This protocol balances the need to explore a significant part of the state space with the need to ensure safety. Additionally, it capitalizes on the fact that at the beginning of the breath exploration is safer and also more valuable. We illustrate this approach in Figure 4: control inputs used in our exploration policy are shown on the top, and the pressure measurements of the ventilator-lung system are shown on the bottom. Precise parameters for our exploration policy are listed in Table 1 in the Appendix.

**Table 1:** Parameters for exploring in the boundary of a PID controller

| $(R, C)$ | $(P, I, D)$ | $(c_{\min}^a, c_{\max}^a)$ | $(t_{\min}^a, t_{\max}^a)$ | $(c_{\min}^b, c_{\max}^b)$ | $(t_{\min}^b, t_{\max}^b)$ | $p_a$ |
| --- | --- | --- | --- | --- | --- | --- |
| (5, 10) | (1, 0.5, 0) | (50, 100) | (0.3, 0.6) | (-20, 40) | (0.1, 0.5) | 0.25 |
| (5, 20) | (1, 3, 0) | (50, 100) | (0.4, 0.8) | (-20, 60) | (0.1, 0.5) | 0.25 |
| (5, 50) | (2, 4, 0) | (75, 100) | (1.0, 1.5) | (-20, 60) | (0.1, 0.5) | 0.25 |
| (20, 10) | (1, 0.5, 0) | (50, 100) | (0.3, 0.6) | (-20, 40) | (0.1, 0.5) | 0.25 |
| (20, 20) | (0, 3, 0) | (30, 60) | (0.5, 1.0) | (-20, 40) | (0.1, 0.5) | 0.25 |
| (20, 50) | (0, 4, 0) | (70, 100) | (1.0, 1.5) | (-20, 40) | (0.1, 0.5) | 0.25 |

**Table 2:** *P* and *I* coefficients that give the best L1 controller performance relative to the target waveform averaged across the six waveforms associated with *PIP* = [10, 15, 20, 25, 30, 35].

| $(R, C)$ | $P$ | $I$ | $D$ |
| --- | --- | --- | --- |
| (5, 10) | 10.0 | 0.2 | 0.0 |
| (5, 20) | 10.0 | 10.0 | 0.0 |
| (5, 50) | 10.0 | 10.0 | 0.0 |
| (20, 10) | 8.0 | 1.0 | 0.0 |
| (20, 20) | 5.0 | 10.0 | 0.0 |
| (20, 50) | 5.0 | 10.0 | 0.0 |

**Figure 4:**
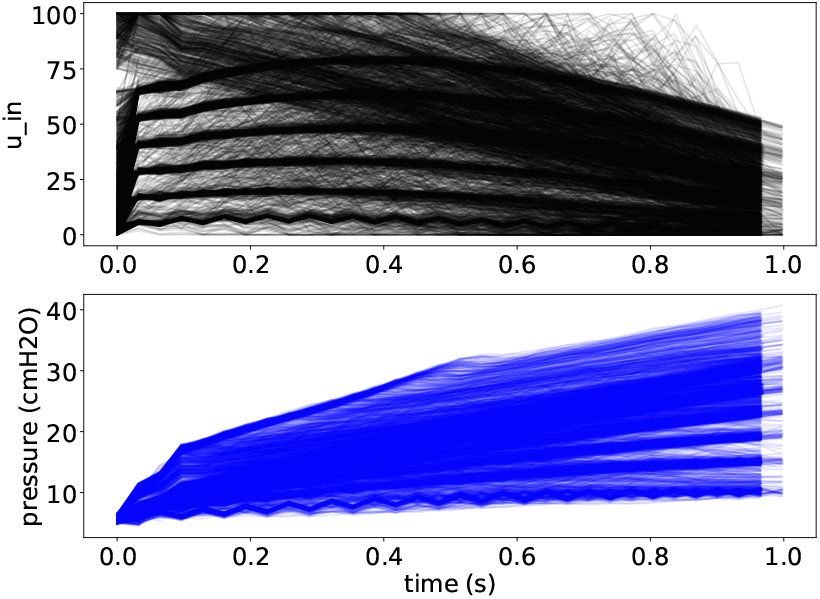
We overlay the controls and pressures from all inspiratory phases in the upper and lower plots, respectively. From this example of the simulator training data (lung setting *R* = 5, *C* = 50), we see that we explore a wide range of control inputs (upper plot), but a more limited “safe” range around the resulting pressures.

### 4.3 Model architecture

Now we describe the empirical details of our data-driven simulator. Due to the inherent differences across lungs, we opt to learn a different simulator for each of the tasks, which we can wrap into a single meta-simulator through code that selects the appropriate model based on a user’s input of *R* and *C* parameters.

The simulator aims to learn the dynamics of the inhalation phase. To more carefully deal with the different behaviors between the “rise” and “stabilize” phases of an inhalation, we learn a collection of models for the very beginning of the inhalation/episode and a general model for the rest of the inhalation, mirroring our choice of exploration policies. This proves to be very helpful as the dynamics at the very beginning of an inhalation are transient, and also extremely important to get right due to downstream effects. Concretely, our final model stitches together a list of *N* = 5 boundary models and a general model, whose training tasks and architectures are described in the paragraphs below (details and minor variations found in Appendix B.2, Table 5).

#### Training Task(s)

We define the boundary training task *i* to be that of predicting the next pressure *p*_*i*+1_, based on the past *i* controls *u*_1_, …, *u*_*i*_ and pressures *p*_1_, …, *p*_*i*_. We define the general training task to be that of predicting the next pressure *p*^*t*+1^, based on the past *H*_*c*_ controls *u*_*t*_*—H*_*c*_, …, *u*_*t*_ and past *H*_*p*_ pressures *p*_*t*_*−*_*H*_^*p*^, …, *p*_*t*_. We featurize the collected data with *H*_*p*_ = 5, *H*_*c*_ = 3 so that this task becomes explicit in the data fed to our neural network model.

#### Boundary Model architecture

Each boundary model is a simple 2-layer feed-forward neural network with 100 hidden units and tanh activation. We trained all models with SGD and L1 loss for 800 epochs. We used batch size 16, learning rate 10^*−*3^ and weight decay 10^*−*3^.

#### General Model architecture

For the general model, we use a 4 layer self-normalizing neural network Klambauer et al. (2017), where each hidden layer has width 100. We trained this model with Adam Kingma & Ba (2017) and L1 loss for 500 epochs. We used batch size 512, learning rate 10^*−*3^ and no weight decay.

#### Evaluation

We evaluate our simulators by holding out 10% of data collected according to our exploration protocol described in 4.2 and computing the average open-loop distance over the inspiratory phases, as described in 4.1. We choose to only focus on the open-loop distance since it is a more reliable description of transfer, minimizing hard-to-analyze interactions between the policy and the simulator. This is also a harder task because it allows us to make use of the random exploration data, rather than restrict us to a specific policy parametrization. In Figure 3, we see a qualitative representation of our simulator’s performance on the *R* = 5, *C* = 50 task. The associated score is 0.30 average open-loop distance normalized by episode length. We provide the plots for the remaining 8 settings in Table 4 and the associated scores in Table 3 (both in Appendix B.1).

**Table 3:** Mean absolute error for the open-loop test under each lung setting.

| (R, C) | Open-loop Average MAE |
| --- | --- |
| (5, 10) | 0.42 |
| (5, 20) | 0.27 |
| (5, 50) | 0.30 |
| (20, 10) | 0.59 |
| (20, 20) | 0.72 |
| (20, 50) | 0.62 |

**Table 4:**
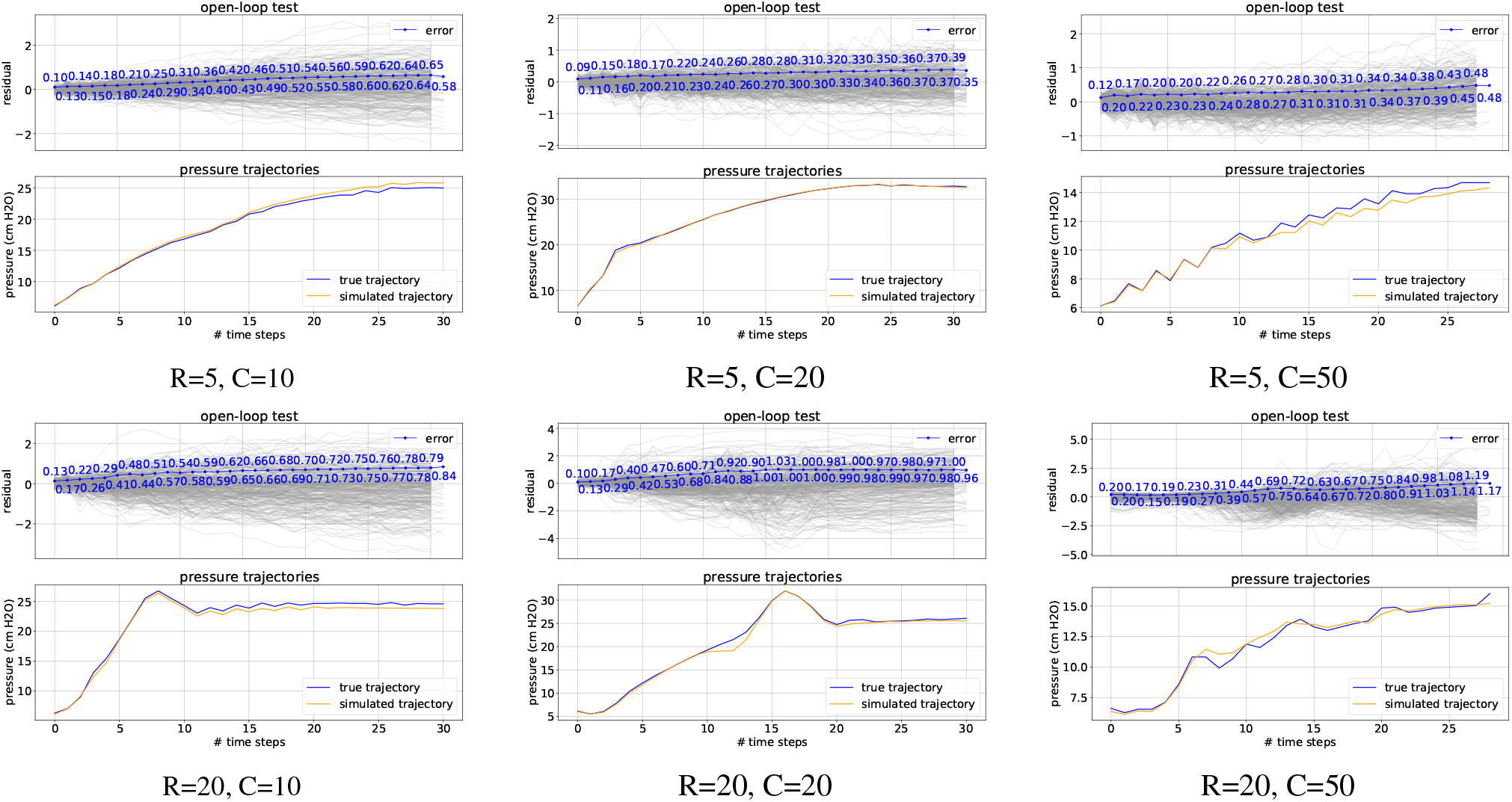
We plot both open-loop testing and pressure trajectories for each of the six simulators corresponding to the six lung settings under consideration. These plots are described more in Section 4

**Table 5:** The primary architectural and training parameters we used for each lung setting.

| (R, C) | # boundary models | weight decay | initial learning rate |
| --- | --- | --- | --- |
| (5, 10) | 5 | 0 | 1e-3 |
| (5, 20) | 5 | 0 | 1e-3 |
| (5, 50) | 5 | 0 | 1e-3 |
| (20, 10) | 5 | 0 | 1e-3 |
| (20, 20) | 5 | 0 | 1e-3 |
| (20, 50) | 15 | 1e-6 | 1e-4 |

## 5 Learning a controller from learned physics

Our experimental results henceforth show that we succeed in the following two tasks:

1. **Tracking improvement:** improve performance for tracking the desired pressure in ISO-specified benchmarks.
2. **Generalization:** improve performance using a **single** deep neural network controller over several lung settings. The controller should improve upon PID even when the PID controllers are trained separately over the different lung parameters.

We first specify the details of the experiments, and then present the results.

### Controller architecture

Our controller is comprised of a PID baseline upon which we learn a deep network correction. We use this general architecture to construct controllers for both the tracking improvement task and the generalization task.

### Baseline PID

The baseline PID of a controller for the tracking improvement task is set to the most performant PID controller from a grid search on the corresponding physical lung setting^2^. The baseline PID of a controller for the generalization task is set to the PID with the lowest average error across a certain subset of lung settings. We describe more details in Appendix A.2 and Table 2.

### Deep Component

On top of the baseline PID, we train a 2-layer neural network correction. The network takes as input a length *k* = 10 history of previous errors *{ϵ*_*t*_ … *ϵ*_*t−k*_*}*, where 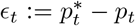, thereby generalizing the class of linear error-feedback controllers (which contains PID). Our network first performs the following feature transformation

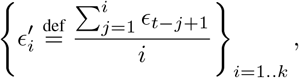

allowing it to react to the error feedback signal averaged over all time scales (further generalizing the PID controller). This is followed by a convolutional layer with kernel width 3 and 5 output channels and finally a fully connected layer.

### Training Objectives

We formalize the training objectives for the two tasks mentioned at the beginning of this section:

1. Tracking improvement: minimize the combined *L*_1_ deviation from the target inhalation behavior across all target pressures on the simulator corresponding to a single lung setting of interest.
2. Generalization: minimize the combined *L*_1_ deviation from the target inhalation behavior across all target pressures *and* across the simulators corresponding to *several* lung settings of interest.

### Training Specifics

The deep component is trained with the Adam optimizer (Kingma & Ba, 2017) via automatic differentiation *through the learned dynamics simulator(s)* with respect to the controller’s parameters. Note that this is *not* possible on a physical lung, and is a key advantage of our methodology. For the learning rate, we start from a large learning rate and decay at plateau until convergence. We detail our choice of hyperparameters in the Appendix.

### 5.1 Experiments

For our experiments, we use the physical test lung to run both our proposed controllers, which were trained on simulators, and the PID controllers that perform best on the physical lung.

To make comparisons, we compute a score for each controller on a given test lung setting (e.g., *R* = 5, *C* = 50) by averaging the *L*_1_ deviation from a target pressure waveform for all inspiratory phases after the initial breath, and then averaging these average *L*_1_ errors over six waveforms specified in ISO 80601-2-80:2018. We choose *L*_1_ so as not to over-penalize breaths that fall short of their target pressures and to avoid engineering a new metric (e.g., with some asymmetry).

We determine the best performing PID controller for a given setting by running exhaustive grid searches over reasonable *P* and *I* coefficients for each lung setting (details for both our score and the grid searches can be found in the Appendix).

#### Tracking improvement

For a given test lung setting, we compare the best performing PID controller for that setting to the controller trained using the simulator for that lung setting. For example, for *R* = 5, *C* = 50, we find the PID controller that performs best and compare it to our proposed controller trained on the simulator for *R* = 5, *C* = 50. We make this comparison on *R* = *{*5, 20*} × C* = *{*10, 20, 50*}*.

#### Generalization

For a *set* of test lung settings, we compare the PID controller with the highest average score over that set of lung settings to a controller that was trained on the simulators corresponding to the same lung settings. We then evaluate performance for both controllers on each lung in the set. We focus on the settings *R* = 20, *C* = [10, 20, 50] since this set is the most commonly seen during the COVID-19 pandemic (Haudebourg et al., 2020).

#### Numerical Results

We benchmark the performance of the proposed controller in the graphs above. As shown in Figure 5, for the tracking improvement task, our separately-trained controllers outperform the best PID controllers tuned specifically to each lung setting. We improve upon the best PID for all lung settings where *R* = 5 and *R* = 20. For the generalization task over *R* = 20 settings, we show that our proposed controller outperforms the best PID controller found over the three *R* = 20 settings in each individual setting. What is more, our controller even outperforms the best PID controller in each setting as well.

**Figure 5:**
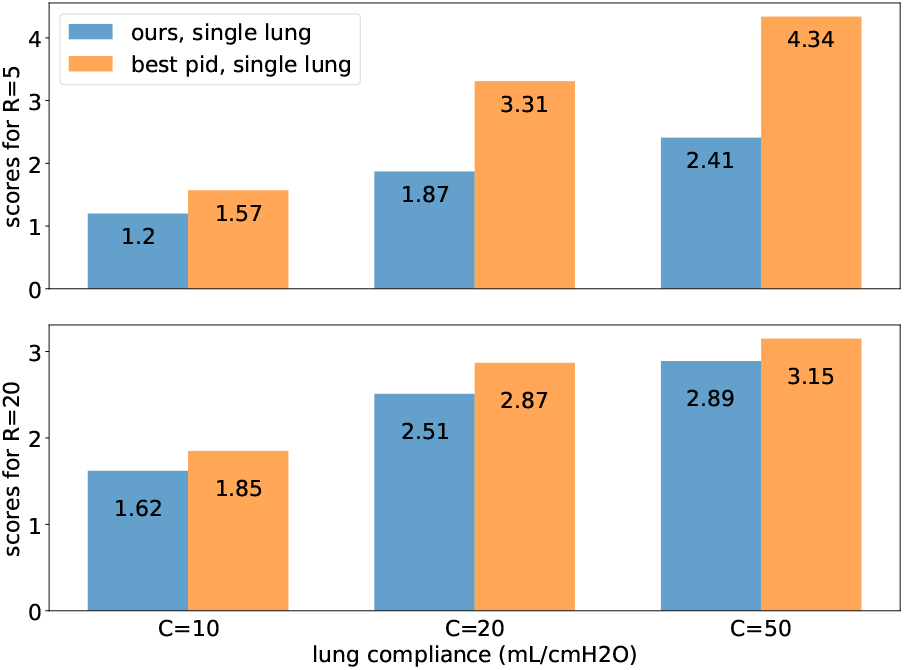
We show that for each lung setting, the controller we trained on the simulator for that setting outperforms the best-performing PID controller found on the physical test lung.

**Figure 6:**
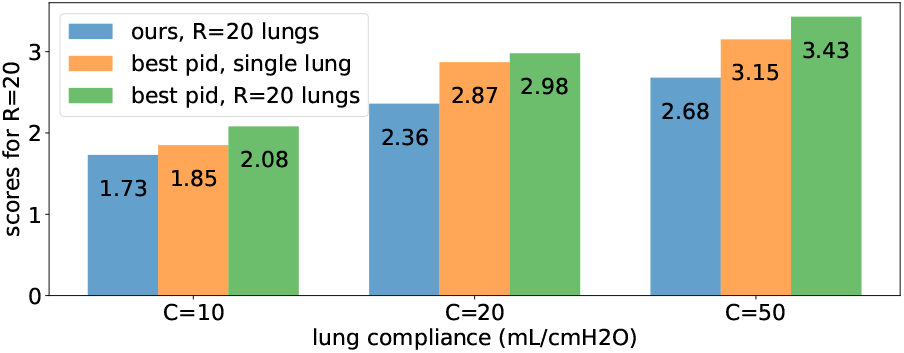
The controller we trained on all three *R* = 20 simulators outperforms the best PID found over the three *R* = 20 settings on the physical test lung. Of note, our controller *also* outperforms the best PID found on each individual setting.

#### Qualitative Behavior

In Figure 7, we can see that the PID controller (orange) is insufficiently expressive to match the target waveform, undershooting the target waveform (dotted line) for most of the breath before overshooting, while ringing the entire time. By contrast, our controller rises quickly to the target pressure and maintains an even pressure for the majority of the inspiratory phase.

**Figure 7:**
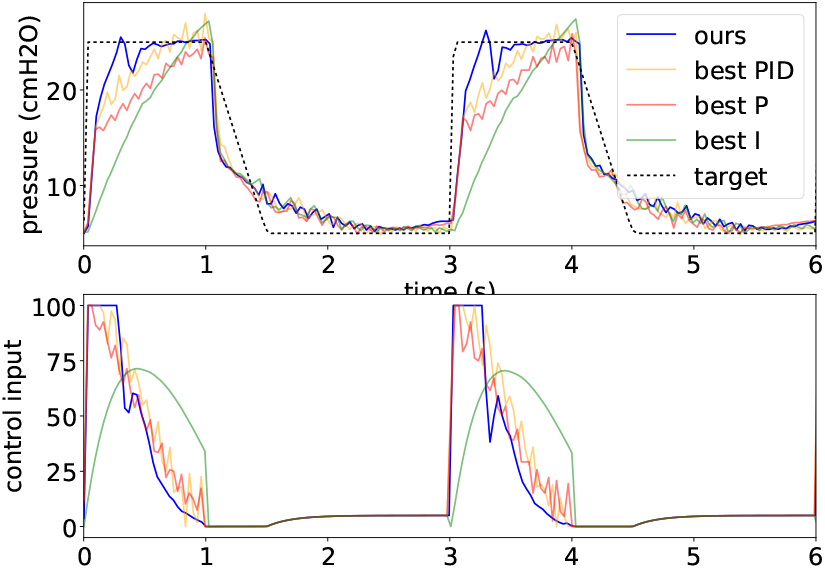
As an example, we compare our method (learned controller on learned simulator) to the best P-only, I-only, and PID controllers relative to a target waveform (dotted line). Whereas our controller rises quickly and stays very near the target waveform, the other controllers take significantly longer to rise, overshoot, and, in the case of P-only and PID, ring the entire inspiratory phase.

## 6 Conclusions and future work

We have presented a machine learning approach to ventilator control, demonstrating the potential of end-to-end learned controllers by obtaining improvements over industry standard baselines. Our main conclusions are

1. The nonlinear dynamical system of lung-ventilator can be modeled by a neural network more accurately than previously studied physics-based models.
2. Controllers based on deep neural networks can outperform PID controllers across multiple clinical settings (waveforms).
3. Deep neural controllers can generalize better than PID controllers across patient lungs characteristics, despite having significantly more parameters.

There remain a number of areas to explore, mostly motivated by medical need. The lung settings we examined are by no means representative of all lung characteristics (e.g., neonatal, child, non-sedated) and lung characteristics are not static over time; a patient may improve or worsen, or begin coughing. Ventilator costs also drive much further research. As an example, inexpensive valves have less consistent behavior and longer reaction times, which exacerbate bad PID behavior (e.g., overshooting, ringing), yet are crucial to bringing down costs and expanding access. Learned controllers that adapt to these deficiencies may obviate the need for such trade-offs.

As part of our study we make our simulators and code available to the scientific community, to lower the entrance cost into this important medical application.

## Data Availability

To be made available shortly

## A Data collection

### A.1 PID residual exploration

The following table describes the settings for determining policies (*a*) and (*b*) for collecting simulator training data as described in Section 4.

### A.2 PID grid search

For each lung setting, we run a grid of settings over *P* and *I* (with values [0.0, 0.1, 0.2, 0.3, 0.4, 0.5, 0.6, 0.7, 0.8, 0.9, 1.0, 2.0, 3.0, 4.0, 5.0, 6.0, 7.0, 8.0, 9.0, 10.0] each). For each grid point, we target six different waveforms (with identical PEEP and breaths per minute, but varying PIP over [10, 15, 20, 25, 30, 35] cmH2O. This gives us 2,400 trajectories for each lung setting. We determine a score for the run by averaging the L1 loss between the actual and target pressures, ignoring the first breath. Each run lasts 300 time steps (approximately 9 seconds, or three breaths), which we have found to give sufficiently consistent results compared with a longer run.

Of note, some of our coefficients reach our maximum grid setting (i.e., 10.0). We explored going beyond 10 but found that performance actually degrades quickly since a quickly rising pressure is offset by subsequent overshooting and/or ringing.

## B Simulator details

### B.1 Evaluation

#### Open-loop test

To validate a simulator’s performance, we hold out 20% of the P and I controller data (480 of the 2,400 trajectories from the PID grid search; see Section A.2). We run the exact sequence of controls derived from the lung execution on the simulator. We define the point-wise error to be the absolute value of the distance between the pressure observed on the real lung and the corresponding output of the simulator, i.e. 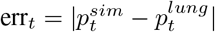. We assess the MAE loss corresponding to the errors accumulated across all test trajectories.

#### Trajectory comparison

In addition to the open-loop test, we compare the true trajectories to simulated ones as described in Section 4.

### B.2 Training Details

For each of the six lung settings, we maintain the same general architecture described in 4.3. We vary some parameters slightly and also have minor variations in training procedure, which we outline in Table 5. Of note, we use a simple learning rate schedule that starts with an initial learning rate and drops the learning rate by a factor of 10 if the loss does not improve after a number of epochs.

## C Controller details

### C.1 Training hyperparameters

We use an initial learning rate of 10^*−*1^ and weight decay 10^*−*5^ over 30 epochs.

### C.2 Training a controller across multiple simulators

To the generalization task, we train controllers across multiple simulators corresponding to the lung settings (*R* = 20, *C* = [10, 20, 50] in our case). For each target waveform (there are six, one for each PIP in [10, 15, 20, 25, 30, 35] cmH2O) and each simulator, we train the controller round-robin (i.e., one after another sequentially) once per epoch. We zero out the gradients between each epoch.

## Footnotes

2 When a sufficient baseline does not exists, we choose a baseline of 0.

